# The Mediterranean Diet Scale (MDS): translation and validation of the Arabic version

**DOI:** 10.1101/2023.06.08.23291163

**Authors:** Raghdah Aljehani, Ghaidaa Aljehani, Hanaa Alharazi, Paula M Horta, Camila Kümmel Duarte, Gabriela Lima de Melo Ghisi.

**Author notes:** **Corresponding author:** Department of Physical Therapy, University of Toronto, Toronto, Canada. 160-500 University Ave, Toronto, Canada M5G 1V7 x. 5261. **Author Contributions:** Conceptualization, RA and GLMG; Methodology, GLMG; Formal Analysis, GLMG; Investigation, RA, GA, HA, PMH, CK; Resources, RA; Data Curation, RA, GLMG; Writing –Original Draft Preparation, GLMG; Writing – Review & Editing RA, GA, HA, PMH, CK; Project Administration, GLMG.

## Abstract

**Objectives:** The self-administered version of the Mediterranean Diet Scale (MDS) has been developed to test the inherent characteristics of this dietary pattern in a quick and simple way, due to the need of this assessment in the clinical and research setting. This study aimed to translate and psychometrically validate the self-administered MDS in Arabic (CRBS-A).

**Methods:** The original (English) version was originally translated to Arabic, followed by back-translation. Next, 10 healthcare providers, followed by 10 cardiovascular disease (CVD) patients rated the face and content validity (CV) of materials, providing input to improve cross-cultural applicability. Then, 200 patients from Saudi Arabia completed the questionnaire, of which factor structure, internal consistency, criterion and construct validity were assessed.

**Results:** Content and face validity was supported based on experts and patients’ reviews (ranges: CV scores 0.9-1.0/1.0 and clarity 3.5 to 4.5/5). Minor edits were made. Subsequent factor analysis revealed 4 factors consistent with the original version of the instrument, all internally consistent. Total CRBS-A α was 0.74. Criterion validity was confirmed by the significantly higher scores in patients who participated in CR. Construct validity was also established by significant associations between MDS scores and monthly family income, having the diagnosis of acute coronary syndrome or with a history of valve repair or replacement, being obese or having dyslipidemia.

**Conclusions:** Overall, these results confirm the validity and reliability of the MDS in Arabic-speaking patients.

## Introduction

Cardiovascular diseases (CVDs) are the most notable causes of disease burden across many nations [1], including Arabic-speaking countries [2,3]. In addition to health impacts, CVDs also carry a substantial economic burden on patients, their families, and societies due to medical costs, diminished productivity from disability and premature death [4-6]. Cardiovascular associations - regionally and internationally – recommend secondary prevention interventions to address this burden, which rely on successful health behaviour change [7-10]. These recommendations are based on findings linking CVDs to modifiable risk factors, particularly unhealthy behaviours such as poor-quality diet [10-12].

Cardiac rehabilitation (CR) is a comprehensive outpatient chronic disease management program designed to facilitate *heart*-health behaviours [9]. CR typically includes core components that address CVD risk factors, including exercise training, patient education, psychosocial management risk factor modification and nutritional counseling [13]. Research has shown that when all these multifactorial components are delivered, CR participation is associated with *reductions of mortality up to 25*%, as well as beneficial effects on morbidity, symptoms, exercise tolerance and capacity, lipid and blood pressure levels, blood pressure, psychosocial functioning and behaviour change [14-16]. In order to tailor adequate CR strategies and sustainable recommendations, it is crucial to fully understand patients’ habits that determine cardiovascular health [17]. One of those is an adequate nutrition [18].

Over the years, various dietary patterns have been studied with the goal to identify the one that is mostly effective on heart health. The Mediterranean Dietary Pattern (MDP) has been greatly studied, with evidence showing its effects on the prevention and treatment of CVDs, as well as on the reduction of CVD mortality (MDP) [19-21]. While the MDP is widely recommended to reduce CVD risk, there is not robust evidence for adherence in CR after the many changes that happened in programs due to the COVID-19 pandemic [22,23].

In this context, questionnaires are considered useful means to collect important information that can support choices related to the care of patients [24]; however, not always their application can be implemented into clinical practices. For example, full-length food frequency questionnaires (FFQ) or food records used in dietary assessments have limitations related to accuracy and time [25], leading to a significant burden on the respondents and healthcare team [26]. The 14-item Mediterranean Diet Score (MDS) was developed to assess adherence to the MDP [27], due to its effectiveness for primary and secondary prevention of CVDs [28-30]. In order to eliminate barriers for its use, a self-administered version of the MDS was developed and validated by Ghisi et al (2018) [31].

The prevalence and control of CVDs is considered the biggest public health challenge in Arabic-speaking countries [32], with risk factors such as obesity more than doubling in the last years [33]. Changes in food consumption patterns have been observed in these countries, including a diet with high content of fat, sugars, sodium and cholesterol [34,35]. In addition, availability of CR programs in Arab countries is extremely limited, with only 8 programs identified in a previous global survey of CR programs [36]. In this context, having a validated questionnaire to evaluate the MDP adherence in Arabic-speaking patients is timely, supporting practitioners to develop and deliver dietary recommendations that are aligned with patients’ needs. Therefore, this study aimed to translate and psychometrically validate the Arabic version of the MDS.

## Methods

### Design and Procedures

The study was reviewed and approved by the King Abdullah Medical City, Saudi Arabia (22-944). Data was collected between September 2022 and January 2023. Permission to conduct this Arabic translation and validation was obtained, and the first author of the original MDS self-administered developmental study was invited to be part of the Arabic project. Authors had no access to information that could identify individual participants during or after data collection.

Following best practices [37], this study included the following steps: 1) translation from English to Arabic and cultural adaptation of the MDS, 2) review by an expert panel and Arabic-speaking patients, and 3) cross-sectional survey for evaluation of psychometric properties.

As the first step, the initial conceptual translation of the scale from English to the target language (Arabic) was performed by an independent translator. Then, a second translator (now blinded to the objectives of the study) back translated the scale. All versions – original, translated, and back translated – were reviewed and combined by the research team into one version, which was reviewed by an expert team comprised of 10 healthcare providers that care for CVD patients (step 2). This expert team assessed face and content validity; the later by rating each item on a 5-point Likert-type scale ranging from 1 = completely irrelevant to 5 = very relevant. Content validity index (CVI) for the items (I-CVI) and scale (S-CVI) were computed. For both, only values higher than 0.80 were considered appropriate [38]. The scale was then revised based on CVI scores and suggestions made by experts. As part of phase 2, a group of 10 individuals from the target population assessed clarity of the scale usinf a 5-point Likert-type scale ranging from 1 = not clear to 5 = very clear. Results were analyzed to further revise the scale.

Finally, psychometric properties were evaluated (step 3). The translated and culturally adapted tool was administered to a convenience sample of cardiac patients from a cardiology center in Saudi Arabia. Patients were invited to participate in the study while attending regular clinical consultations. Those that accepted, signed the consent form, and completed the Arabic MDS in-person. The following psychometric properties were assessed following the Consensus-based Standards for the selection of health Measurement Instruments (COSMIN) taxonomy [39]: factor structure, internal consistency, construct, and criterion validity.

### The self-administered version of the MDS

The original 14-item MDS was developed in Spanish [27] and later translated to English [40]. The English version was used to develop the self-administered version [31]. This version included pictures to facilitate comprehension of questions and a simplified language. It is comprised of 13 items with yes/no options. The items are divided into 4 domains as follows: meats and processed foods; olive oil and sauce; fruits, vegetable, nuts, and legumes; and, fish and seafood [31]. Each item is scored as 0 or 1 in accordance with MDP adherence [41]. The final score ranges from 0 to 13 (< 5 = low adherence to MDP; > 10 = high adherence to MDP) [31]. The self-administered version of the MDS is also available in Brazilian [42] and Chinese [43] languages.

### Participants

The inclusion criteria for patient participants were confirmed cardiac diagnosis or presence of cardiovascular risk factors, 18 years of age or older, and being able and willing to provide informed consent. The exclusion criteria for patient participants were inability to eat by mouth or having food and dietary restrictions (e.g., nut allergies) and having any impairment which preclude the participant’s ability to answer the questionnaire (e.g., visual). A minimum of 200 participants was required following the recommendation for factor analysis [39].

Patient participants self-reported their sociodemographic and clinical characteristics, including the following: age, sex, cardiovascular diagnosis and risk factors, living area, educational level, and family income. In addition, CR participation (yes/no) was self-reported to assess criterion validity.

### Data Analysis

First, exploratory factor analysis was performed, using the main component method for factor extraction. Only factors with eigenvalues > 1.0 were considered. Item factor loadings > 0.3 were used in finalizing the items for each factor and interpreting the factors [44]. Next, internal consistency was determined by calculating Cronbach’s *alpha* values of the scale and subscales. Cronbach’s *alpha* higher than 0.70 was considered acceptable [45].

Criterion validity was assessed by differences in MDS total scores by CR participation using independent samples *t*-tests. Construct validity was assessed by exploring the associations between self-reported sociodemographic characteristics of participants and MDS scores using Pearson’s correlation, independent samples *t*-tests, and analysis of variance.

Finally, a descriptive analysis of the Arabic MDS was performed. The Statistical Package for Social Sciences v. 28 (SPSS Inc., Chicago, IL, USA) was used for data analysis. The level of significance for all tests was set at 0.05.

## Results

### Translation, cultural adaptation and review by an expert panel and patients

Following translation and back translation, a revised Arabic MDS version was reviewed by an expert panel (n=10). The I-CVI ranged from 0.9 and 1.0 and the S-CVI was 0.9. These results identify an acceptable content validity for the Arabic MDS. For patients’ review (n=10), clarity scores ranged from 3.5 to 4.5/5 (mean 4.2±0.3). Given these results, two items were culturally adapted as follows: pork removed from items 10 and 11 due to religious and cultural reasons. In addition, items 6 and 13 were rephrased to increase clarity. Appendix 1 displays the Arabic MDS.

### Cross-sectional survey for evaluation of psychometric properties

Overall, 200 patients completed the Arabic MDS. Their sociodemographic and clinical characteristics are presented in Table 1.

**Table 1:** Characteristics of patient participants and corresponding mean total MDS scores (N=200)

| **Characteristic** |  | **MDS Total Score**<br>**mean±SD** | **p\*** |
| --- | --- | --- | --- |
| *Age*, mean±SD | 46.5±13.6 | - | - |
| *Sex*, n (%) |  |  | 0.11 |
| Male | 92 (46.0) | 8.3±2.3 |  |
| Female | 108 (54.0) | 7.7±2.4 |  |
| *Area of Living*, n (%) |  |  | 0.51 |
| Urban | 180 (90.0) | 7.9±2.4 |  |
| Rural | 19 (9.5) | 8.5±1.7 |  |
| *Highest educational level*, n (%) |  |  | 0.91 |
| Illiterate | 9 (4.5) | 7.7±2.7 |  |
| Less than High School | 5 (2.5) | 8.2±1.9 |  |
| High School | 62 (31.0) | 7.8±2.5 |  |
| College certificate or diploma | 109 (54.5) | 8.0±2.3 |  |
| Higher education | 15 (7.5) | 8.3±2.0 |  |
| *Monthly family income*, n (%) |  |  | 0.02 |
| Under SAR\$10,000 | 78 (39.0) | 6.6±2.5 | |
| Between SAR\$10,001-20,000 | 76 (38.0) | 7.6±2.2 | |
| Between SAR\$20,001-\$30,000 | 38 (19.0) | 7.5±2.4 | |
| Higher than SAR\$30,001 | 8 (4.0) | 9.1±2.2 | |
| *Cardiac Diagnosis/Procedures, n (%)* |  |  |  |
| Percutaneous Coronary Intervention | 24 (12.0) | 8.1±2.2 | 0.72 |
| Arrhythmia | 22 (11.0) | 7.4±2.6 | 0.25 |
| Acute Coronary Syndrome | 21 (10.5) | 9.0±2.2 | 0.02 |
| Acute Myocardial Infarction | 16 (8.0) | 8.0±2.3 | 0.90 |
| Valve disorders | 16 (8.0) | 8.5±3.3 | 0.40 |
| Cardiac Device | 11 (5.5) | 8.1±2.2 | 0.85 |
| Valve Repair/Replacement | 9 (4.5) | 9.4±3.2 | 0.03 |
| Heart Failure | 8 (4.0) | 7.4±1.6 | 0.50 |
| Coronary Artery Bypass Graft | 2 (1.0) | 7.5±2.1 | 0.78 |
| *Risk factors, n (%)* |  |  |  |
| Family History of CVD | 79 (39.5) | 7.7±2.4 | 0.30 |
| Sedentary/inactivity | 72 (36.0) | 7.8±2.5 | 0.50 |
| Smoking | 64 (32.0) | 8.1±2.1 | 0.70 |
| Diabetes Type II | 60 (30.0) | 8.3±2.3 | 0.23 |
| Hypertension | 46 (23.0) | 7.8±2.4 | 0.52 |
| Obesity | 44 (22.0) | 5.7±2.3 | 0.02 |
| Depression | 43 (21.5) | 8.2±2.5 | 0.44 |
| Dyslipidemia | 34 (17.0) | 6.5±2.3 | 0.03 |
| *CR participation, yes (%)* |  |  | 0.002 |
| Yes | 22 (11.0) | 9.5±2.2 |  |
| No | 178 (89.0) | 7.6±2.3 |  |

MDS, Mediterranean Diet Scale; SD standard deviation; CVD cardiovascular diseases; CR cardiac rehabilitation.
\*p is used for association of characteristics and total mean score, tested using t-tests, analysis of variance or Pearson's correlation, as applicable.
Note: Income shown in Saudi Riyal (SAR). 1 SAR corresponds to USD\$0.47 (currency: March 1, 2023)

Results from the Kaiser-Meyer-Olkin index (KMO=0.78) and Bartlett’s Sphericity tests (*X²=*403.921, p<0.001) indicated that the data were suitable for exploratory factor analysis. Four factors were extracted, similar to the original validation [25]. These factors represented 62.3% of the total variance and were all internally consistent (Cronbach’s alpha ranged from 0.70-0.79). Factor 1 reflected items related to meats/processed foods (5 items), factor 2 related to olive oil/sauce items (3 items), factor 3 related fruits/vegetables and nuts/legumes (4 items), and factor 4 related to fish/seafood (1 item). Table 2 presents the factor structure of the Arabic MDS SV, including item loadings. The total Cronbach’s alpha of the scale was 0.74, confirming the internal consistency of the scale.

**Table 2:** Factor loadings from exploratory factor analysis, reliability of factors and number of ‘yes’ responses per item, N=202.

| Item | Factors |  |  |  | Yes, n (%) |
| --- | --- | --- | --- | --- | --- |
|  | 1. Meats and Processed Foods | 2. Olive Oil and Sauces | 3. Fruits, Vegetables, Legumes and Nuts | 4. Fish and Seafood |  |
| 1. Do you use olive oil as the main source of fat when you cook? |  | 0.721 |  |  | 123 (61.5) |
| 2. Do you use at least four tablespoons or more of olive oil when cooking your food each day? |  | 0.733 |  |  | 118 (59.0) |
| 3. Do you eat two servings or more of vegetables each day? |  |  | 0.689 |  | 100 (50.0) |
| 4. Do you eat three servings or more of fruit each day? |  |  | 0.629 |  | 101 (50.5) |
| 5. Do you eat less than one tablespoon of butter, hydrogenated margarine or cream each day? | 0.466 |  |  |  | 111 (55.5) |
| 6. Do you drink less than one serving of sweet or sweetened drinks each day? | 0.408 |  |  |  | 100 (50.0) |
| 7. Do you eat three servings or more of legumes a week? |  |  | 0.469 |  | 137 (68.5) |
| 8. Do you eat three servings or more of fish or seafood each week? |  |  |  | 0.474 | 87 (43.5) |
| 9. Do you eat one serving or more of nuts each week? |  |  | 0.453 |  | 129 (64.5) |
| 10. Do you eat poultry (chicken or turkey) more often than meat (beef, veal, hamburger, or sausage)? | 0.453 |  |  |  | 164 (82.0) |
| 11. Do you limit red meat and processed meats to one serving or less one or two times a week? | 0.452 |  |  |  | 132 (66.0) |
| 12. Do you eat less than three servings of sweets or pastries each week? | 0.405 |  |  |  | 119 (59.5) |
| 13. Do you add some flavorings to your food, such as a mixture of tomatoes, garlic, onions and leeks, two or more times a week? |  | 0.652 |  |  | 171 (85.5) |
| Variance explained | 27.4% | 16.8% | 9.6% | 8.5% | - |
| Eigenvalues | 2.9 | 1.5 | 1.3 | 1.1 | - |
| Reliability | 0.79 | 0.78 | 0.75 | 0.70 | - |

\* Item culturally adapted.

With regard to criterion validity, significant associations were observed between total scores and CR participation. Those who participated in CR had significantly higher adherence to the MDP (i.e., higher MDS scores) than those who did not participate (p=0.002; Table 1).

As also shown in Table 1, with regard to construct validity, there were significant associations between MDS scores and monthly family income (p=0.02), having the diagnosis of acute coronary syndrome (p=0.02), a history of valve repair or replacement (p=0.03), being obese (p=0.02) or having dyslipidemia (p=0.03). In this context, those with a family monthly income under SAR$10,000 presented a significantly lower MDP adherence than their counterparts. In addition, those who had a diagnosis of acute coronary syndrome, those that were obese, with dyslipidemia, or those who underwent a valve repair or replacement, presented MDP adherence than those without the diagnosis or a history of these procedures (Table 1).

### Adherence to the MDP

The total score for the self-administered version of the MDS in this study was 11.0±8.0/13. Overall, 57 (28.5%) patients were classified as having high adherence to MDP and 29 (14.5%) as having low adherence. Table 2 also displays the number of patients that adhere to MDP recommendations (i.e., responded “yes” to each item asked in the questionnaire). Items with the highest adherence included seasoning foods with a combination of tomato, garlic, onions, or leek (n=171; 85.5%), eating poultry more often than red meat (n=164; 82.0%), and eating three servings or more of legumes a week (n=137; 68.5%). One item - eating three servings or more of fish or seafood each week – had the percentage of yes lower than 50% (n=87; 43.5%).

## Discussion

Research has proven the benefits of the MDP for both primary and secondary prevention of CVDs [46]. However, multi-level barriers affect adherence to dietary recommendations, including in participants of CR programs [47]. Having a validated and convenient tool to assess MDP adherence – and ultimately guide clinical and educational practices – is timely in countries like Saudi Arabia where a small percentage of population meet dietary recommendations and programmes to improve dietary behaviours are urgently needed to reduce the current and future burden of disease [48]. Therefore, this study established the Arabic version of the MDS to assess adherence to the MDP through a multi-step process that included translation, cultural adaptation and assessment of psychometric properties. Content and face validity was supported based on experts and patients’ reviews. Subsequent factor analysis revealed 4 factors consistent with the original version of the instrument [31], all internally consistent. Criterion validity was confirmed by the significantly higher scores in patients who participated in CR. Construct validity was also established by significant associations between MDS scores and monthly family income, having the diagnosis of acute coronary syndrome or with a history of valve repair or replacement, being obese or having dyslipidemia. Overall, these results confirm the validity and reliability of the MDS in Arabic-speaking patients.

The self-administered version of the MDS – originally developed in English [31] and later translated and validated to Portuguese [41] and Chinese [42] – was developed to simplify the assessment of MDP adherence in clinical and research settings. This is the 4^th^ language available and other validation projects are underway (including Spanish and French). The Cronbach’s alpha of the Arabic MDS was 0.74, which is higher than previous studies (range: 0.42-0.69). The total MDS score of this score was higher than the Canadian (10.2±1.9) [31], Portuguese (6.94±1.99) [41], and Chinese (7.7±2.4) [42] versions. Items identified in the current study as the ones with the highest adherence included flavouring foods with a combination of tomato, garlic, onions, or leek, eating poultry more often than red meat, and eating three servings or more of legumes a week. These results are similar to responses identified in the previous studies using the MDS; in Canada [31], the items with highest adherence included eating poultry more often than meat and flavouring foods with a combination of tomato, garlic, onions or leek; in Brazil [41], eating three servings or more of legumes a week and flavoring foods with a combination of tomato, garlic, onions, or leeks; in China [42], flavouring foods with a combination of tomato, garlic, onions or leek. On the other hand, the self-reported consumption vegetable in the Arab-speaking sample was lower than the other samples (50% vs. 85% in Canada, 79% in Brazil and 87% in China) [31,41,42], despite the fact that the Saudi dietary guidelines strongly recommend a higher intake of vegetables [49].

As previously mentioned, construct validity of the Arabic MDS was confirmed in this study, as well as in other versions of this scale [31,41,42]. Specifically, those with lower monthly family income had significantly lower MDP adherence than their counterparts. Despite its complexity, the impact of socioeconomic on dietary pattern is undeniably important [50-52]. In general, people with higher income tend to follow the MDP recommendations [53]; however, in any context, recommendations to follow healthy dietary habits should be given to CVD patients. Future research examining the changes in adherence to the MD over time and after educational interventions is warrantied, including in groups of individuals with low socioeconomic status.

Those who participated in CR had significantly higher adherence to the MDP (i.e., higher MDS scores) than those who did not participate. These results support the effects of participation in CR – and delivery of nutrition counselling – on adherence to heart-healthy dietary patterns [54]. Furthermore, studies should describe the strategies, tools, and techniques employed within the interventions used to adhere to the MDP and barriers and facilitators to follow this behaviour.

There are some limitations in this study. First, findings are limited to one center from 1 Arabic-speaking country (Saudi Arabia); results cannot be generalizable to the other 21 countries in the world that speak Arabic. Second, there may be selection and retention bias, which also limits the generalizability of the findings. Third, because of the nature of this cross-sectional study, causal conclusions cannot be drawn; it is recommended to test if the scale is sensitive to change before and after a nutritional or educational intervention. The final limitation pertains to the assessment of other psychometric properties such as test-retest reliability, which should be explored in future studies.

## Conclusion

In conclusion, the Arabic MDS is a valid and reliable tool that can be used to assess MDP adherence of Arabic-speaking cardiac patients. It is hoped that this scale supports the adequate assessment of food intake and changes in dietary habits, which plays a highly significant role in the care of people living with a cardiovascular condition.

## Supplementary Materials

Appendix 1: Arabic MDS

## Data Availability

Data cannot be shared publicly because of Ethics Restrictions. Data are available from the King Abdullah Medical City Institutional Data Access / Ethics Committee (contact via 1st author) for researchers who meet the criteria for access to confidential data.

